# GenTIGS: A database empowering research and clinical insights on rare genetic disorders with an Indian perspective

**DOI:** 10.1101/2025.04.01.25325014

**Authors:** Iliyas Rashid, Pooja S, Shivranjani Moharir, Rakesh Mishra

## Abstract

Rare Genetic Diseases (RGDs) are conditions caused by gene mutations affecting less than 1 in 2,000 individuals, as per World Health Organization (WHO). India, being the most populous country in the world has a high prevalence of these diseases. The situation is worsened further due to the practice of consanguinity in several communities and limited genetic testing. Multiple recent developments, including advancements in genetic sequencing, precision medicine and patient advocacy supported by collaborative networks and government initiatives offer hope for improved diagnosis and treatment. With an objective to bring what is known about these rare disorders on a single platform for the researchers, clinicians and various stakeholders, we have developed a database, GenTIGS, that is a ‘to go platform’ for information about these diseases. GenTIGS is a comprehensive database, that includes the information about the genes and pathogenic variants, clinical symptoms for in-depth exploration of RGDs with a focus on globally reported disorders, especially those prevalent in India. For facilitating ease of use, GenTIGS encompasses an array of features and data points crucial for researchers and clinicians delving into RGD domains, ensuring efficient information retrieval. This data delivery system provides information on 2315 RGDs and 2779 associated genes, including 707 globally reported disorders prevalent in India. It also includes details on 3525 clinical symptoms and 307340 pathogenic variants for these disorders. GenTIGS provides extensive data, comprehensive range of analytical tools and resources for researchers, clinicians, and academicians, facilitating in-depth exploration of genes and variants associated with rare genetic disorders and features supporting advancements in genetic medicine by enhancing understanding and analysis within the scientific and clinical community.

Accessible: https://db.tigs.res.in/gentigs/

## Introduction

Rare Genetic Diseases (RGDs), stemming from gene mutations or function-altering variations, present unique challenges due to their infrequent occurrence and consequential impact on affected individuals (1,2). Monogenic RGDs are known to exhibit various inheritance patterns— be it dominant or recessive and can be autosomal or sex-linked (3). Often chronic and severely disabling, these conditions impose a substantial burden on healthcare systems (4,5). The World Health Organization estimates that over 6,000 rare diseases affect 1 in 2,000 individuals, particularly in pediatric conditions, (6,7,8) accounting for 3.5% to 5.9% of the global population, with some diseases being rare within specific demographic groups (9,10). The realm of rare diseases historically lacked attention in scientific enquiry due to limited resources and technology. However, with the emergence of advanced techniques like Next-Generation Sequencing (NGS), the landscape of research has shifted, leading to the identification and documentation of new rare diseases and conditions (11). While progress has been made in certain disorders, the field remains in its early stages (12), with challenges in identifying affected families and securing adequate DNA samples for analysis (13). The International Rare Diseases Research Consortium (IRDiRC) Perspective highlights that many disease genes and variants remain undiscovered, emphasizing the need to accelerate discovery efforts (14).

India’s high prevalence of rare genetic disorders, estimated to affect 70 million individuals (15), is exacerbated by consanguineous marriages, lack of awareness, and limited access to genetic testing (16). This burden is compounded by social stigma, financial constraints, and inadequate healthcare infrastructure (17). Ongoing research initiatives in India aim to understand the genetic basis of rare disorders, develop novel diagnostics, and explore innovative treatments, facilitated by collaborative networks, clinical trials, and advanced genetic sequencing technologies (18,19). The promise of precision medicine, gene therapy, and personalized treatment strategies offers hope for enhancing outcomes and quality of life for those with rare genetic conditions (20,21). Patient advocacy groups, non-profit organizations, and government initiatives in India collaboratively raise awareness, provide support services, and advocate for individuals with rare genetic disorders, addressing challenges through research promotion, policy changes, and public awareness efforts (22,23).

Several global platforms dedicated to rare genetic disorders offer specialized resources that enable users to access, analyze, and interpret data across various aspects of diagnosis, treatment, and research. OMIM, for example, covers human genetic disorders and offers detailed information on clinical features, inheritance patterns, genetic mutations, associated genes, and related research (24). Similarly, Orphanet (25), GARD (26), GRDR (27), and NORD (28) support researchers, healthcare professionals, and advocacy groups by facilitating collaboration and providing essential information to improve the lives of individuals with rare genetic disorders (Table 1). In addition, NCBI resources (29) offers valuable genetic data on gene-disease associations, genetic testing options, and clinical variant interpretations. Furthermore, platforms like Human Phenotype Ontology (30), gnomAD (31), MalaCard (32), and the Monarch Initiative (33) collectively enhance rare disease research by fostering collaboration, standardizing data, interpreting genetic variations, and integrating comprehensive clinical and genetic information to accelerate diagnosis and treatment.

**Table 1.** Full names of abbreviated resources useful for analyzing genetic disorders.

| **Resource Abbreviation** | **Full Resource Name** | **Access Link** |
| --- | --- | --- |
| ClinGen | Clinical Genome Resource | https://clinicalgenome.org/ |
| ClinVAR | Clinical Variation Database | https://www.ncbi.nlm.nih.gov/clinvar/ |
| EMBL-EBI OLS | European Bioinformatics Institute Ontology Lookup Service | https://www.ebi.ac.uk/ols4 |
| GARD | Genetic and Rare Disease Information Center | https://rarediseases.info.nih.gov/ |
| gnomAD | Genome Aggregation Database | https://gnomad.broadinstitute.org/ |
| GRDR | Global Rare Diseases Patient Registry Data Repository | https://grdr.hms.harvard.edu |
| GTR | Genetic Testing Registry | https://www.ncbi.nlm.nih.gov/gtr/ |
| GUaRDIAN | Genomics for Understanding Rare Diseases: India Alliance Network | https://guardian.genomes.in/ |
| HPO | Human Phenotype Ontology | https://hpo.jax.org/ |
| IRDIRC | International Rare Diseases Research Consortium | https://irdirc.org/ |
| MedGen | Medical Genetics Database | https://www.ncbi.nlm.nih.gov/medgen/ |
| NCBI | National Center for Biotechnology Information | https://www.ncbi.nlm.nih.gov/ |
| NIDAN Kendra | National Inherited Disorders Administration Kendras | https://dbtindia.gov.in/dbt-press/inauguration-nidan-kendras-and-ummid-launch-dbt-website |
| NORD | National Organization for Rare Disorders | https://rarediseases.org/rare-diseases/ |
| NPRD | National Policy for Rare Diseases | https://rarediseases.mohfw.gov.in/ |
| OMIM | Online Mendelian Inheritance in Man | https://www.omim.org/ |
| UMMID | Unique Methods of Management of Inherited Disorders | https://dbtindia.gov.in/scientific-directorates/information-systems-biology/ummid |

Numerous initiatives have been undertaken in India to address rare genetic disorders, focusing on offering services and data management related to genetic variations and rare conditions prevalent in the Indian population. These efforts aim to support research, diagnosis, and treatment strategies (34–36). Genome-wide data from over 2,800 individuals across 260 South Asian groups revealed unique founder events leading to recessive diseases, highlighting the potential to reduce disease burden through the identification of population-specific, disease-associated genes (37). India’s health priorities have shifted in recent years, with increasing focus on non-communicable diseases (NCDs), which accounted for 61.8% of all deaths in 2016. There has been a considerable development in the area of rare disease research in India recently (38). The GUaRDIAN consortium (2019) in India brings together clinicians and researchers to study rare genetic disorders, utilizing advanced technologies and animal models (39). The Indian government’s initiatives, including NPRD 2021, UMMID, and NIDAN Kendras, (Table 1) focus on reducing rare disease incidence through prevention, early detection, treatment, genetic counseling, and specialized care to improve healthcare access for patients in India (40). Despite the availability of global patient care databases and services, India has made limited progress in establishing a comprehensive gene repository for rare genetic diseases. Such a repository is crucial for integrating data and studying inheritance patterns, which could significantly improve early diagnosis, treatment, and research. Achieving this requires collaboration among healthcare institutions, researchers, and policymakers. A well-organized gene repository would streamline data collection, foster research collaboration, and enhance information sharing, thereby enabling the identification of genetic patterns and the prioritization of resources for improved genetic counseling and treatment guidelines.

In India where access to basic healthcare is a significant challenge over 72 million people are affected by rare diseases (41) further straining an already burdened healthcare system (42). In this context, an initiative such as GenTIGS supports clinical research by consolidating diverse information on a single platform to enhance understanding and treatment of prevalent rare genetic disorders. GenTIGS bridges gaps in rare genetic disorder knowledge and patient care by exploring the complexities, challenges, and initiatives in the field. It provides a strategic vision to enhance understanding, diagnosis, and management of these conditions. With global resources dedicated to rare genetic diseases, we aim to improve research, patient care, and the effectiveness of interventions. This platform offers key information on disease prevalence, symptoms, ongoing research, and guidance to support those affected by rare genetic diseases.

## Materials and Methods

## 1. Database Creation

The database development, data curation, and query management were executed using Linux, Apache, MySQL, PHP, and Perl (LAMPP). A relational database management system MySQL was utilized to structure tables for diverse data management at the backend, running on the Linux operating platform. The database was designed with multiple tables and applied bidirectional normalization to rows and columns to eliminate redundancies. Additionally, indexing was implemented across several tables to optimize query retrieval times.

## 2. Collection, curation, and population of data in the database tables

We utilized NCBI resources (29) as the primary data source for compiling and integrating information on RGDs. Our approach involved gathering comprehensive data on RGDs and their associated genes through an exhaustive search. Following the initial data collection, we enhanced our dataset by incorporating additional curated information from various reliable sources. Data collection was initiated by compiling a list of rare genetic disorders (RGDs) and their associated gene names from case reports and research articles using PubMed (29). In the next process, Indian prevalent disorders were segregated from the broader list based on case reports and research articles specifically reflecting disorders reported in India. To further enrich the information of the listed RGDs, we sourced responsible variants for each disorder from ClinVAR (43) and ClinGen (44). To enhance the depth of information, we also gathered inheritance modes, disorder categories, clinical symptoms, aliases, and relevant annotations from OMIM (45), NCBI-MedGen (46), GARD (26) and GTR (47). Detailed gene information, sequences, and associated annotations were retrieved from NCBI-Gene (48) and human genome GRCh38.p14 (49). MalaCards (32) was also used to obtain supplementary data, disorder subcategories and classification, and integrated with our listed RGDs. A dataflow diagram illustrates the collection and curation of datasets from various resources for integration and management in the database, providing different webpages for browsing and analysis (Fig. 1). All datasets and annotations were curated according to the schema of the respective databases and managed accordingly by writing Perl Programs. Several in-house Perl programs were developed using the DBI (Database Interface) module for database connection, schema design, table definition, and data management. The individual program addressed specific tasks, including data curation, mapping, comparison and integration of relevant datasets obtained from various sources, and populating them into respective tables. Additionally, Perl scripts such as “NCBIGeneDB_Mapper.pl and GeneSeqParse.pl” (50), were employed for parsing gene information and sequences and managing them. Finally, GenTIGS data delivery system was incorporated with efficient data browsing and analysis facilities.

**Figure 1.**
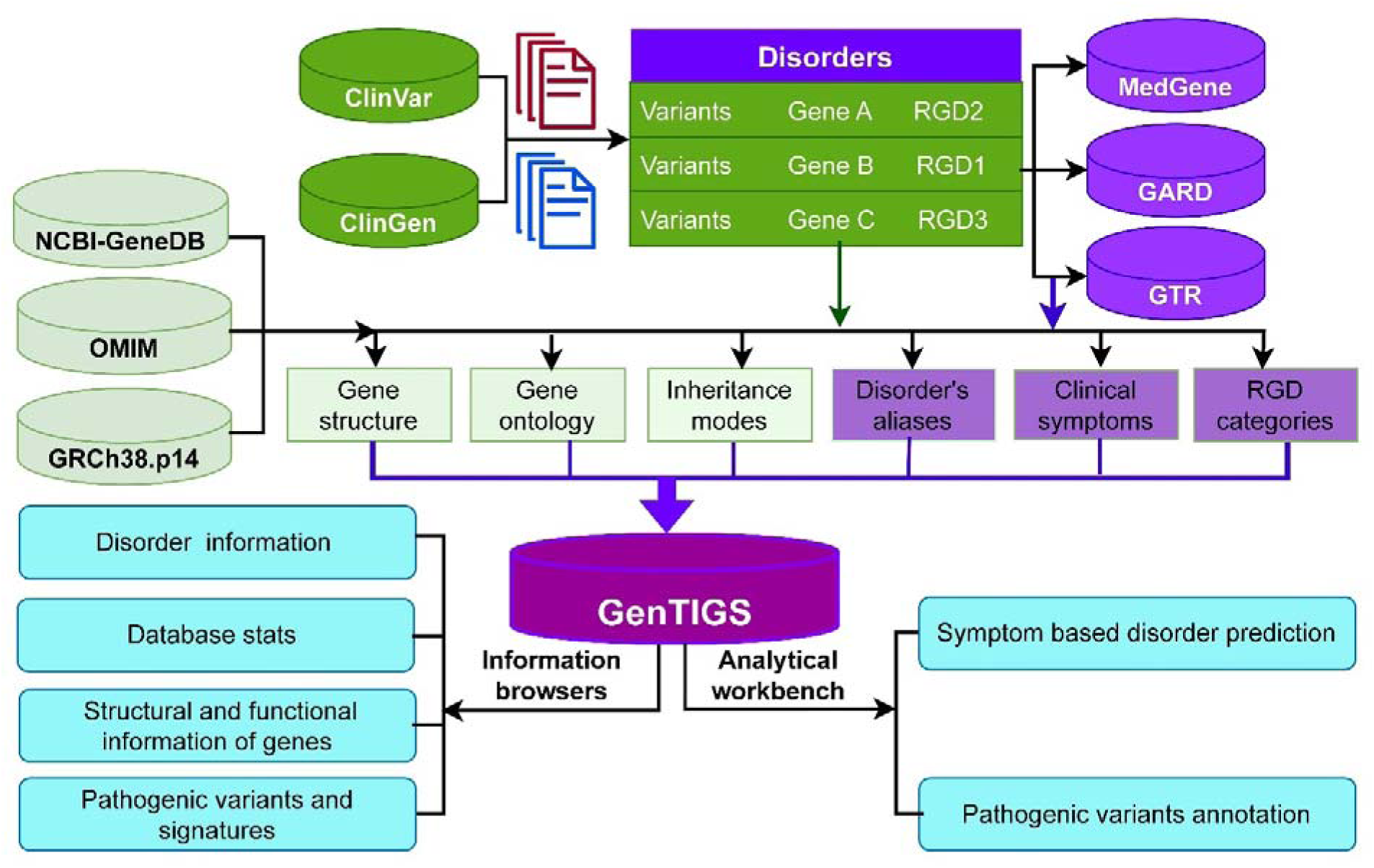
Architecture and data flow diagram of GenTIGS. The schematic outlines the processes of collecting, segregating, enriching, and curating information on rare genetic disorders from multiple data sources, with the final output accessible via user-friendly web interfaces for browsing and analysis.

## 3. Metadata analysis

Variant Call Format (VCF) files from the IndiGenomes project, which includes 1,029 self-reported healthy males and females over the age of 30 from diverse regions across India (51), were made publicly available. These files were subsequently downloaded and processed using a custom script, ‘IndiGenMapper.pl,’ to identify pathogenic variants associated with autosomal dominant and recessive inheritance patterns in this cohort of self-declared healthy individuals. This script aligns variants from VCF files with ClinVar data in the GenTIGS database, focusing on genes associated with disorders prevalent in India. The custom script categorized mapped variants as ‘identical’ (exact match of mutational positions and alleles) or ‘similar’ (same mutation site, different genotypes) exampled in table 2. The mapped variants were further classified into ‘pathogenic with conflicts’ and ‘pathogenic without conflicts’ and organized into the database. The Venny tool (52) was used to visualize the frequencies of identical and similar variants, categorized as pathogenic or nonpathogenic. In this analysis, variants reported in the GRCh38.p14 genome from ClinVar data were considered for annotation, as IndiGenomes used the same reference genome strain.

**Table 2.** Representation of Identical and Similar Mapped Variants. Presenting a detailed comparison of clinically significant variants identified in two datasets: IndiGen and ClinVar. Variants are classified as identical or similar based on their genomic positions, allele changes, and corresponding clinical data.

| Example of Identical mapped Variants | Similar mapped |
| --- | --- |
| • IndiGen: chr13:51944170:C>T<br>• ClinVar: chr13:51944170:C>T<br>• ClinVar Entry:<br>NM_000053.4(ATP7B):c.3182G>A<br>(p.Gly1061Glu)<br>• Condition: Wilson disease (AR)<br>• Exact match of mutational positions and alleles | • IndiGen: chr10:89247612:C>T<br>• ClinVar: chr10:89247612:AC>A<br>• ClinVar Entry:<br>NM_000235.4(LIPA):c.37del (p.Val13fs)<br>• Condition: Wolman disease (AR)<br>• Same mutation site, different genotypes |

## 4. Web Interface

For web-based information delivery and analysis, user interactive dynamic web pages were designed and implemented using LAMPP, CGI (common gateway interface), Ajax, JavaScripts, CSS (cascading style sheets) and HTML technologies.

## Results

This database is exclusively dedicated to storing information on rare genetic disorders and their associated genes, as reported globally, along with a segregated list of disorder prevalence in India based on case reports. The GenTIGS web interface offers user-friendly menu items such as ‘Home,’ ‘Disorders’ and ‘Analysis’. Apart from these diseases specific menus, it also has ‘contact’, ‘feedback’ and ‘keyword search’ options that further facilitate the ease of use and make the interface more interactive. The web-based workbench of GenTIGS includes the ‘Disorders’ and ‘Analysis’ menu items for retrieving, browsing, and analyzing gene information associated with an RGD. Continuous updates are implemented as new information emerges. It serves as a user-friendly search and data delivery system, simplifying data retrieval and facilitating data analysis of monogenic disorders.

## 1. Home menu

The homepage provides an overview of the database and displays statistics with the latest updates. It gives information about the purpose served by the database, how and from where the content is curated, and the significance of the database. GenTIGS currently includes 2315 globally reported RGDs associated with 2779 genes, of which 707 RGDs are prevalent in India across 30 major disorder categories linked to 3525 clinical conditions and influenced by 307340 pathogenic variants (Fig. 2). It also includes information on aliases, inheritance modes, and subcategories of RGDs. A detailed breakdown of the total subcategories and RGD-associated genes across various categories, along with their association with single or multiple genes, is provided in table 3.

**Figure 2.**
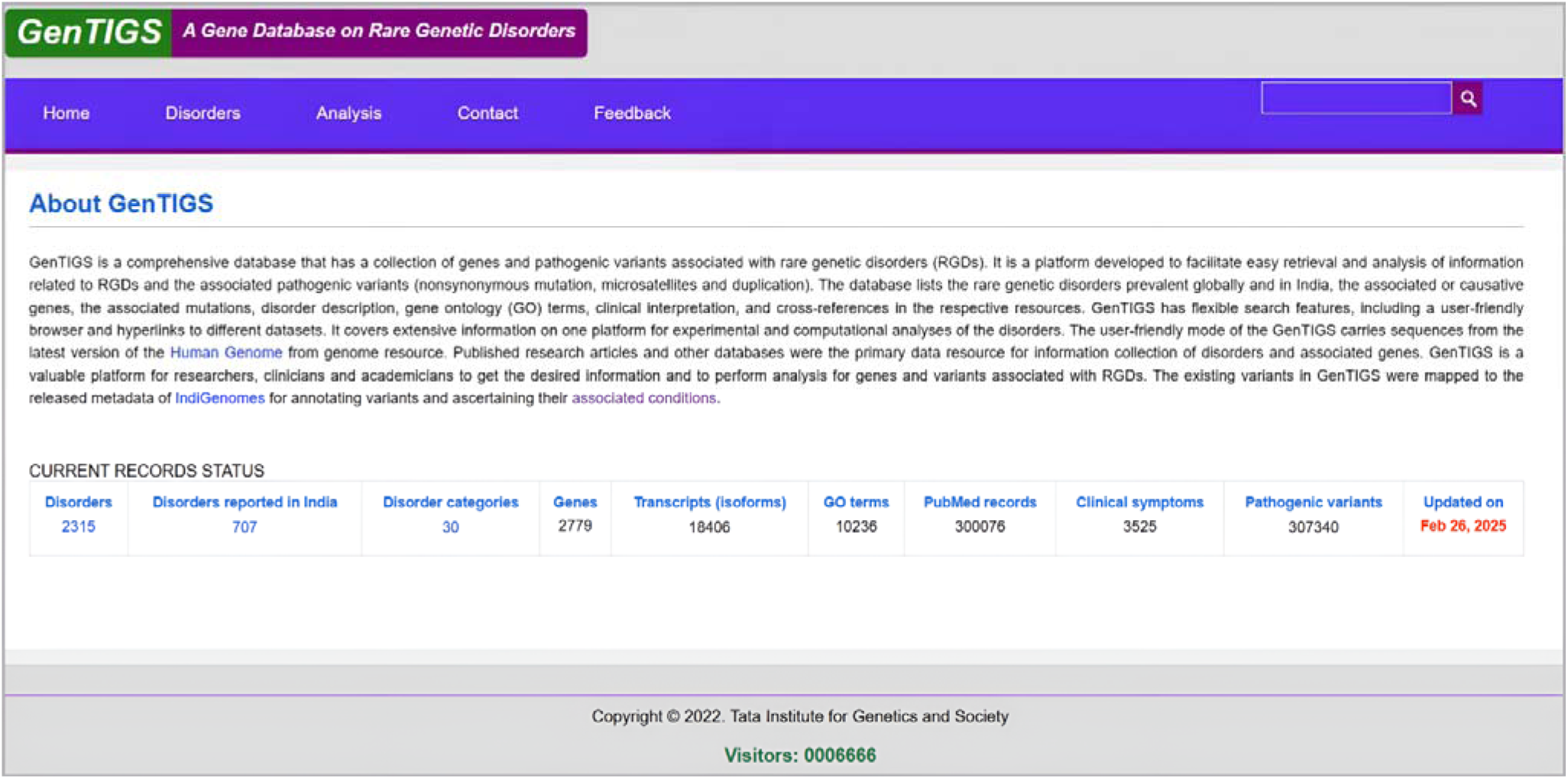
GenTIGS Data Delivery System. Displays global and India-specific disorder prevalence, associated genes, and clinical conditions.

**Table 3.** Major disorder categories. Summary of subcategories, total RGDs, and single-gene versus multi-genes associated disorders across major disorder categories.

| Category name | Total Subcategory | Total RGD | RGD Associated Genes |  |
| --- | --- | --- | --- | --- |
|  |  |  | Single Genes | Multiple Genes |
| Aging disorders | 0 | 2 | 2 | 0 |
| Blood disorders | 12 | 34 | 18 | 16 |
| Bone disorders | 16 | 93 | 63 | 30 |
| Cancer disorders | 25 | 40 | 16 | 24 |
| Cardiovascular disorders | 10 | 14 | 4 | 10 |
| Developmental /Cancer | 0 | 1 | 0 | 1 |
| Developmental/Multisystemic disorders | 0 | 3 | 2 | 1 |
| Ear disorders | 1 | 2 | 0 | 2 |
| Ear disorders/Hair disorders | 0 | 1 | 1 | 0 |
| Endocrine disorders | 8 | 18 | 12 | 6 |
| Eye disorders | 9 | 26 | 14 | 12 |
| Eye disorders/Ear disorders | 1 | 3 | 0 | 3 |
| Gastrointestinal disorders | 8 | 11 | 5 | 6 |
| Hair disorders | 0 | 2 | 1 | 1 |
| Immune disorders | 12 | 36 | 25 | 11 |
| Liver disorders | 1 | 3 | 1 | 2 |
| Metabolic disorders | 27 | 118 | 82 | 36 |
| Metabolic disorders/Lysosomal storage disorders | 10 | 31 | 21 | 10 |
| Multisystemic disorders | 1 | 18 | 12 | 6 |
| Nephrological disorders | 9 | 21 | 7 | 14 |
| Neurodegenerative disorders | 22 | 50 | 36 | 14 |
| Neurodevelopmental disorders | 12 | 36 | 19 | 17 |
| Neuromuscular disorders | 11 | 57 | 36 | 21 |
| Neuronal disorders | 3 | 7 | 6 | 1 |
| Oral disorders | 3 | 8 | 7 | 1 |
| Pain syndrome | 0 | 1 | 0 | 1 |
| Reproductive disorders | 2 | 2 | 1 | 1 |
| Respiratory disorders | 8 | 11 | 4 | 7 |
| Skin disorders | 25 | 55 | 39 | 16 |
| Tumor/Cancer | 0 | 3 | 1 | 2 |
| **30** | **236** | **707** | **435** | **272** |

## 2. Disorders menu

The “Disorders” menu in GenTIGS offers two flexible dropdown options: (i) ‘Disorders Reported in India’, which focuses on data specific to India, and (ii) ‘Worldwide Disorders’, which provides a global perspective. These options allow for efficient access to RGD information, catering to both regional and global disorder data on RGDs.

### 2.1. Disorders reported in India

The “Disorders Reported in India” section functions as the RGD summary webpage, serving as the central information hub within GenTIGS for those disorders that are reported in India. It consolidates and presents key data in a summarized, tabular format, offering users a clear and accessible overview of disorder of interest. At the top of the ‘RGD Summary’ page, a dropdown menu lets users select an RGD of interest that is prevalent in India, displaying detailed information for the chosen RGD. The retrieved content includes the RGD and its aliases, major disorder categories, Mendelian inheritance patterns, and associated genes. The inheritance mode and disorder category, displayed beneath the RGD name, are hyperlinked to their respective web pages for more detailed information. Additionally, the disorder summary webpage provides detailed descriptions of the associated gene(s), including the gene name, a brief overview, chromosomal information, cytogenetic band, genomic accession, gene location, and the number of exons reported for each gene. A hyperlinked column within the table connects users to various external gene databases—such as OMIM, gnomAD Browser, Ensembl, GeneCards, and ClinGen—allowing easy navigation and access to cross-database gene information. A ‘More’ hyperlink in the gene information table leads to the ‘gene details’ page, which reveals information on mRNA, transcript variants, and isoforms. These entries are linked to PubMed IDs referencing articles where each isoform has been reported. Another table in ‘gene details’ displays the variants identified in the same gene as part of the IndiGenome project—a large-scale genomic initiative from India, which includes 1,029 genome samples from adult males and females across the nation. Additionally, the gene details page features another table representing gene ontology (GO) annotations.

The ‘RGD Summary’ page under “Disorders Reported in India” features a central table displaying variant information, showing the total number of variants identified in genes associated with the selected disorder. This table includes a selection list of clinically significant variants, including pathogenic variants, which enables users to explore information of selected clinically significant variants reported in genes linked to the chosen RGD. The ‘Disorders Reported in India’ section includes a detailed table at the bottom, which presents various reported clinical symptoms, medical terminology, descriptions, and the occurrence frequencies associated with selected RGD. Additionally, there is an interactive selection list that allows users to filter and display symptoms based on the frequency of their occurrence for a specific RGD.

In addition to the detailed information, each disorder is linked to multiple widely used databases, such as Orphanet, OMIM, GARD, GTR, MedGen, MalaCards, EMBL-EBI OLS, Monarch Initiative, Human Phenotype Ontology, and NORD. These links facilitate GenTIGS data validation and provide global access to comprehensive datasets from each database, offering diverse and specialized information.

### 2.2. Globally Reported Disorders

The “Worldwide Disorders” is accessible under the “Disorders” menu in GenTIGS. It provides information on disorders and their associated genes reported from various regions around the world, including India. While offering a global perspective, it serves as a superset of the “Disorders Reported in India”. It enables users to easily access an overview of relevant worldwide disorders, including disorder names, associated genes, and cross-references to well-known databases. This resource is valuable for stakeholders such as researchers, clinicians, and patients seeking knowledge on monogenic disorders. Monogenic disorders are primarily caused by mutations in a single gene, but variations within that gene or in multiple genes can contribute to similar traits. These disorders typically follow Mendelian inheritance patterns, even when multiple genes independently influence the phenotype.

## 3. Analyzing information

The ‘Analysis’ menu has tools for symptoms-based disease prediction and metadata analysis that enhance the utility and visibility of the database (Fig. 2). The menu contains two dropdown items: (i) ‘Symptoms-based disorder prediction’ which allows users to explore diseases linked to specific symptoms, and (ii) ‘Metadata Analysis’ offering analysis overview. These features offer seamless access to disorder data and analysis tools, supporting in-depth research and providing a comprehensive compendium of rare genetic variants for better insights into genetic conditions.

### 3.1. Disorder Prediction Interface

The symptoms-based disorder prediction tool is accessible through ‘Analysis’ dropdown menu item in the user interface. A flexible user-friendly query interface is provided, offering a concise description and a manual outlining the steps for initiating disorder predictions. The interface includes a category selection list, which allows users to choose a specific category of interest or to select all categories. Upon selection, the corresponding clinical symptoms associated with the chosen category (or all categories) are displayed in a list on the left-hand side of the selection box. Users can then select one or more clinical symptoms from the list and transfer them to the adjacent box on the right side by clicking the ‘double greater’ sign button (>>).

Once clinical symptoms are selected, users can initiate the disorder prediction process by clicking the “Predict Disorder” button. The predicted disorder(s) and relevant information are then presented dynamically below the query area, in the same interface window, providing an immediate and interactive result display.

### 3.2. Variant annotations

We developed a program that queries metadata from human population genomes to identify pathogenic variants and their related conditions. This tool was applied to the recently released IndiGenomes dataset, that includes genomes sequenced from 1029 individuals. The tool facilitated annotating the variants and associating them to the different conditions reported in GenTIGS. The results of this analysis are accessible via the GenTIGS platform and can be retrieved through the ‘IndiGenomes Metadata’ option in the ‘Analysis’ menu of the user interface. This interface features a dropdown list of various clinical significance for identically mapped variants to retrieve and display information in a centralized table.

During variant annotation and mapping of the IndiGenomes VCF file onto ClinVar in the GenTIGS database for disorders reported in India, two types of mappings were observed. The first involved identical variants, where the position and mutation were the same in both datasets. The second type consisted of similar variants, where the position was identical, but the variant type and mutational base pairs differed. The integration of the IndiGenomes dataset with the GenTIGS database led to the identification of 8,645 variants, including 4,826 identical mutation sites, 3,819 similar mutation sites, and 1,131 pathogenic variants, which encompass both identical and similar mutations (Fig. 3A). Among these, 1,131 distinct pathogenic sites were categorized into 432 identically mapped variants and 699 similar mapped variants (Fig. 3A). A more detailed analysis of these pathogenic variants, categorized by clinical significance, is shown in Fig. 3B. Comprehensive information regarding all 432 identically mapped pathogenic variants, associated with phenotypic conditions and Mendelian inheritance patterns. Every identically mapped variant can be accessed using above described ‘IndiGenomes Metadata’ section under the ‘Analysis’ menu.

**Figure 3.**
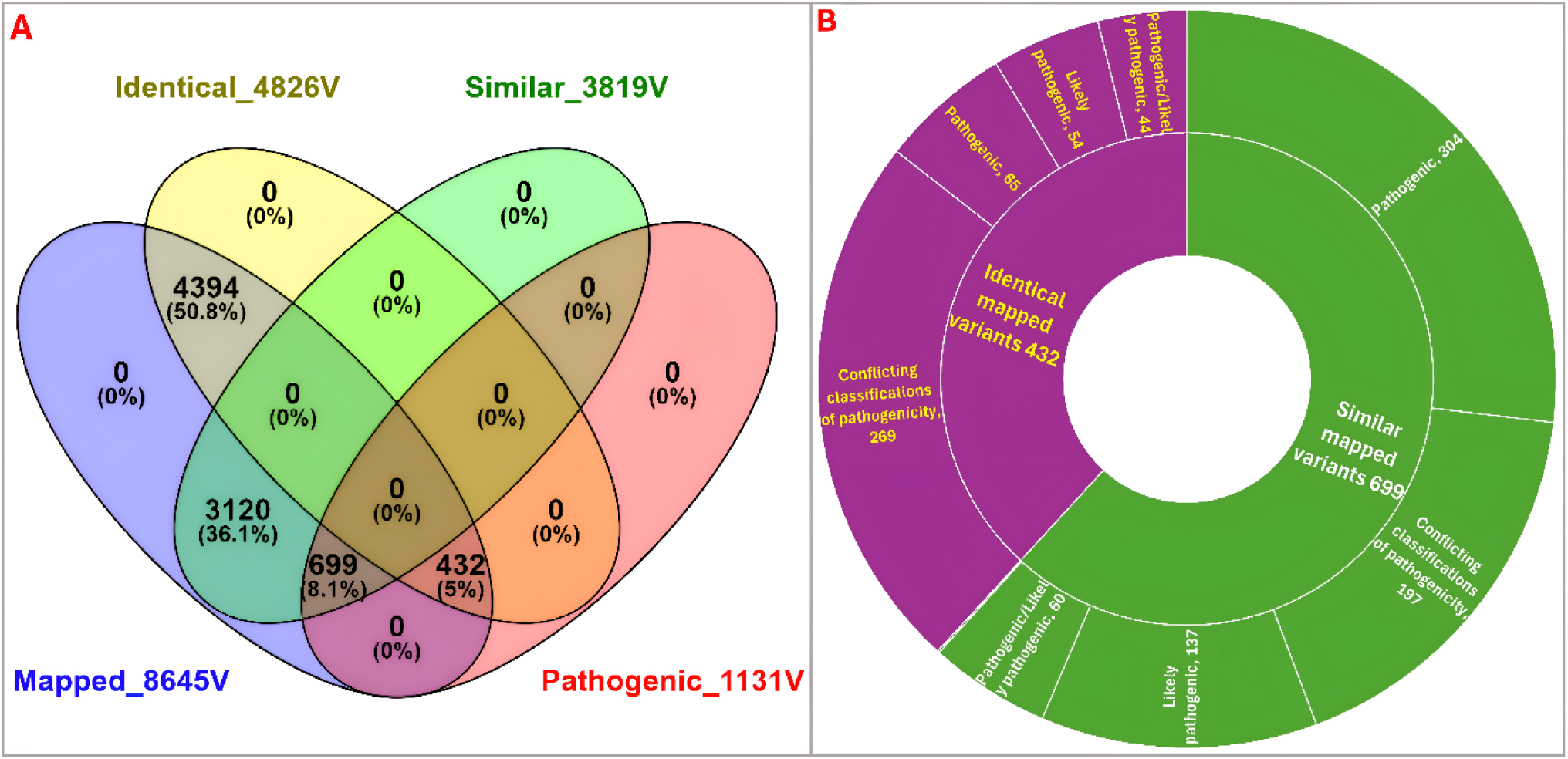
Annotation of IndiGenomes variants. (A) The Venn diagram illustrates the overlap of clinically significant variants in GenTIGS and IndiGenome across four datasets-identical, similar, mapped and pathogenic variant sets. (B) The sunburst chart further classifies the 1131 clinically significant variants into categories: pathogenic, likely pathogenic and variants with conflicting pathogenicity.

An in-depth analysis of the 65 identical pathogenic variants depicted in Fig. 3B identified 14 dominant phenotypes (Table 4) and 37 recessive phenotypes (Table 5) of RGD, all of which have been reported in India. The remaining pathogenic variants were related to X-linked and multi-mode inheritance patterns. Furthermore, all clinically important variants, both pathogenic and benign, were added to the database for reference and analysis. The variants with similar mapping in Fig. 3 are listed with detailed information on their clinical significance in supplementary file ‘SupplementaryData_S1.xlsx’. These findings highlight the importance of precise mapping in identifying exact mutations in clinical genomics.

**Table 4.**
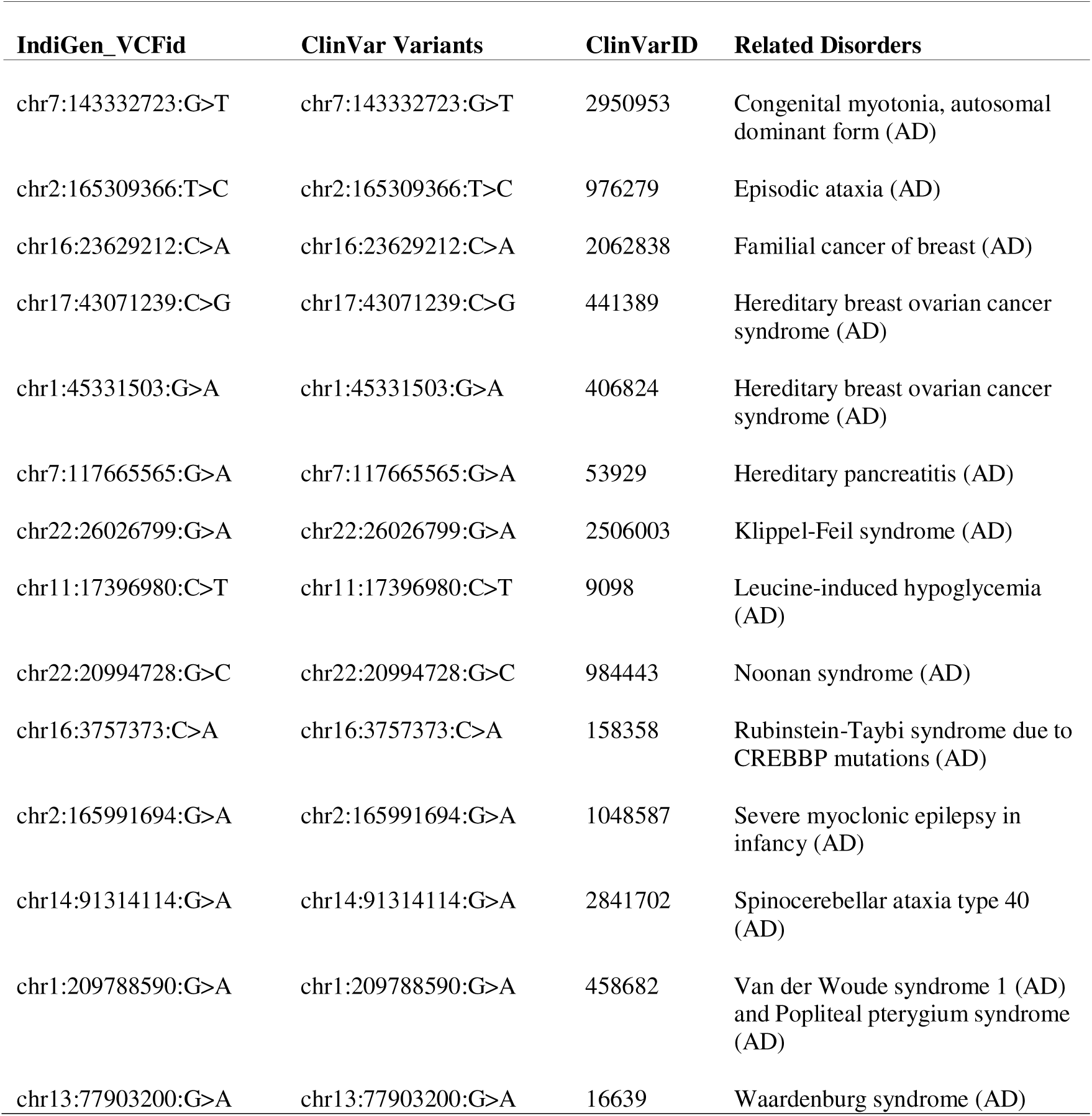
Annotation of variants linked to autosomal dominant disorders. This table lists annotated pathogenic variants from the IndiGenomes VCF along with their association to dominant disorders reported in India through GenTIGS labelling.

**Table 5.**
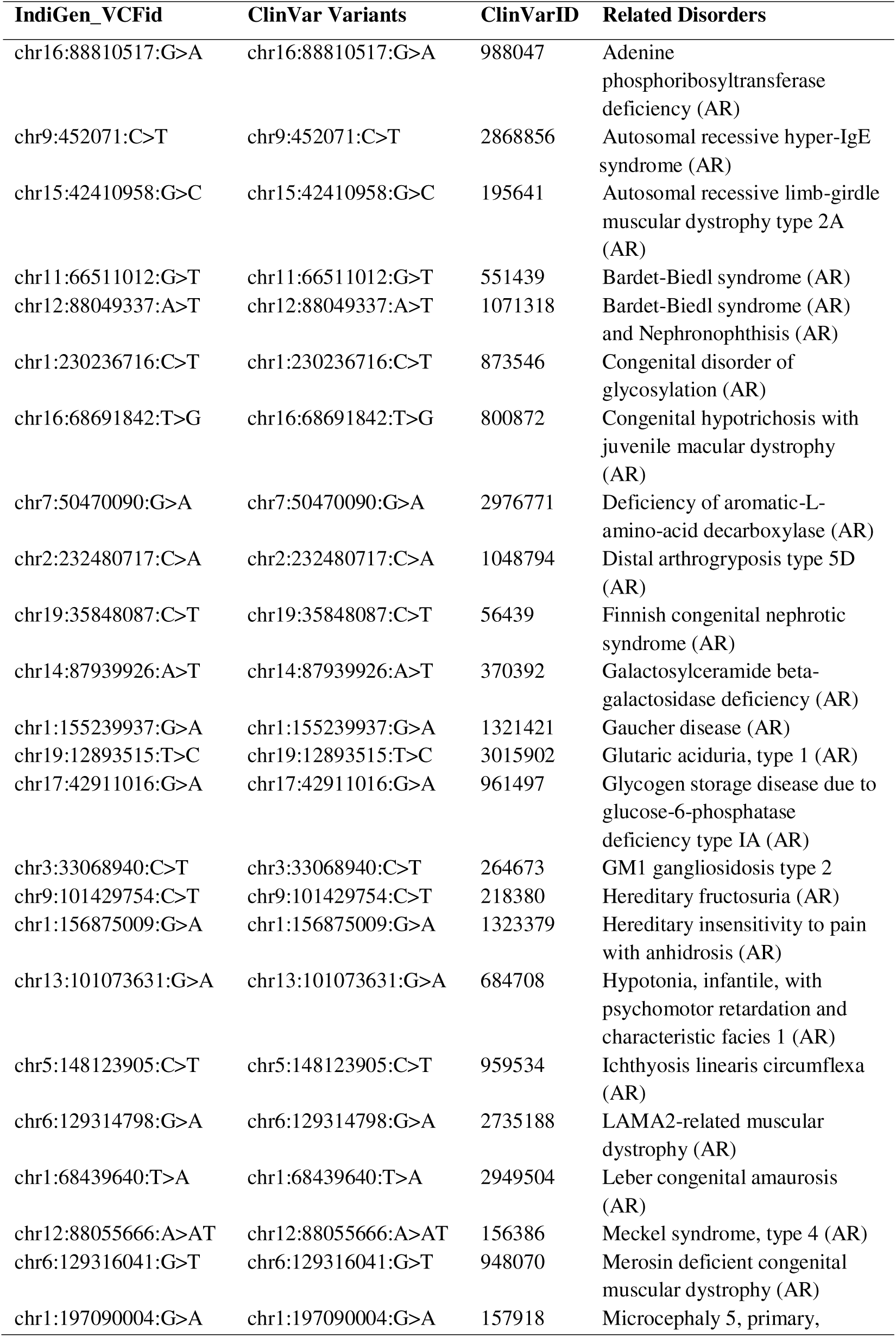
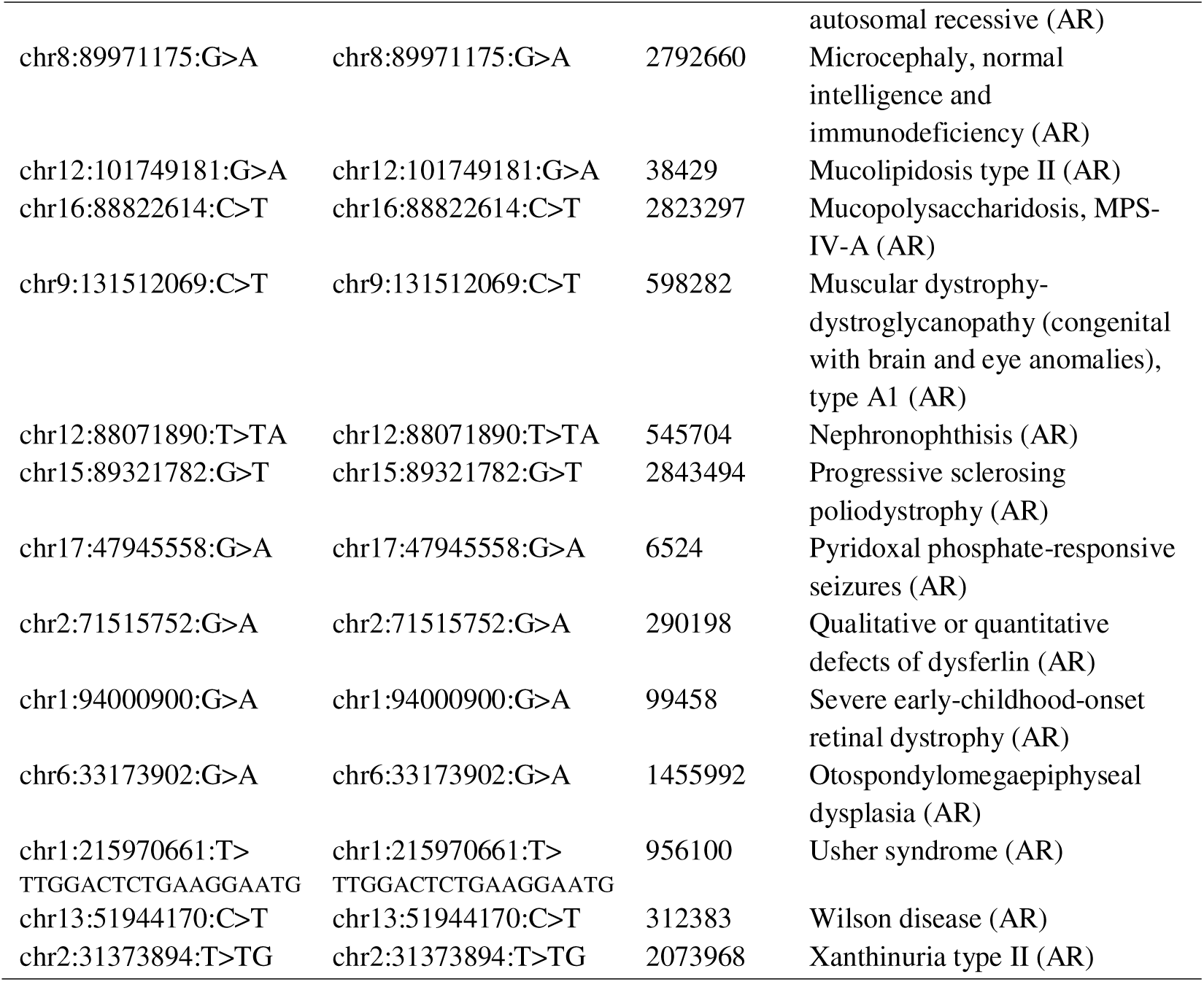
Annotation of variants linked to autosomal recessive disorders. This table lists annotated pathogenic variants from the IndiGenomes VCF along with their association to recessive disorders reported in India through GenTIGS labelling.

## 4. Keyword search

The interface includes a powerful keyword search feature that allows users to input various search terms, such as disorder names, aliases, gene names, gene symbols, and gene IDs, to retrieve relevant records from the database. Depending on the type of keyword entered, the system presents tailored views to display the most pertinent information. These views are designed to ensure that the data is presented in a clear and organized manner, facilitating easy access to the details associated with each keyword. Additionally, the search functionality is optimized to handle large datasets, providing users with fast and accurate results.

## Discussion

The diversity of genetic conditions in India, including the prevalence of inherited genetic diseases across different populations, necessitates the resources that account for regional and ethnic variations (42). As highlighted by India State-Level Disease Burden Initiative Collaborators (2017), India’s vast ethnic diversity and the presence of founder mutations in certain populations underscore the need for a comprehensive genetic resource along with real world data that can cater to these specific needs (51). Such databases provide insights into both the local and global contexts of genetic disorders and their detailed genetic information, bridging the gap in understanding and diagnosis of RGDs in the Indian setting (39,54). By integrating data from major international repositories, alongside region-specific information, this approach is particularly useful for studies and clinical applications for regional as well as global prospectives (55,56). GenTIGS was developed through the integration of multiple data sources, including Indian prevalent RGD data, to improve the comprehensiveness of the database within this context. This facilitates in-depth exploration of RGDs with a global focus, while also addressing the unique genetic landscape of India. Lagunes-García (2020) proposed a method for collecting diverse disease information from public sources and their cross-referencing to create unified resources with multiple datapoints (57). The disorders included in GenTIGS are based on multiple data points gathered from reputable sources. Linking that information to external resources enhances data reliability and allows researchers to cross-validate findings with other databases, thereby ensuring the quality and accuracy of the information. The detailed covering of multiple information on RGDs helps users understand epidemiology and genetics with associated conditions. Importantly, the inclusion of clinical symptoms and their frequencies further enhances the practical utility of the platform for clinicians, enabling better diagnosis and management of RGDs (27). In the context of family information management related to inherited conditions (58), the inclusion of inheritance modes associated with each disorder in GenTIGS allows users to analyze genetic diseases based on their transmission patterns.

The development of enriched web services for biomedical data visualization (59), clinical decision support systems (60), and interactive tools for meta-analysis of diagnostic test accuracy (61) has significantly advanced healthcare by improving data analysis, visualization, and decision-making. Building on these advancements, GenTIGS distinguishes itself with its ability to efficiently search for disorders, genes, aliases, and identifiers, offering refined search parameters that enhance user experience and enable the retrieval of targeted information. The optimized search function ensures rapid and accurate data retrieval, which is essential for both clinical and research applications. Furthermore, the analytical tools for disorder prediction and variant annotation, along with the organized presentation of results, facilitate easy interpretation of data. These features assist clinicians and researchers in identifying relevant genes, understanding clinical conditions, and navigating gene-disease associations and variant details with greater ease and accuracy.

GeneTIGs leverages tailored genetic testing, expanded research, and regional databases to enhance diagnosis, personalized medicine, and genetic counseling, especially in areas affected by founder effects (62). Its “Symptoms-Based Disorder Prediction” feature aids clinicians in predicting potential RGDs from clinical symptoms, streamlining diagnostics. The platform also integrates metadata analysis tools for annotating genetic variants, such as those from the IndiGenomes dataset, improving understanding of genetic diseases in the Indian population. This supports genotype-phenotype correlation studies to refining diagnostic precision and disease comprehension (63,64).

The inclusion of the IndiGenomes dataset, which is derived from over 1000 healthy Indian genomes, provides a unique opportunity to study the genetic diversity of the Indian population and its implications for rare genetic disorders. We utilized GenTIGS to perform a genome wide comparative analysis of the IndiGenomes released dataset. The comparative analysis revealed a set of identically mapped pathogenic variants associated with both autosomal and recessive modes of genetic disorders reported from different parts of the world. Recessive disorder is caused when both the copies of a gene need to carry the associated pathogenic variant for the manifestation of the disease phenotype. If only one copy carries the variant, symptoms typically do not manifest in the clinical conditions, though the individual is a carrier (65,66). A dominant disorder is caused when only one copy of the allele carrying the pathogenic variants (received from either parent) is sufficient for the disorder to develop. These conditions are linked to specific clinical outcomes, which may manifest in infancy or later in life, with varying severity (67,68,45). In the present study, we identified the variants in IndiGenomes dataset, that were already reported in ClinVAR and had an ‘rs’ number. The pathogenic mutations in genes such as MUTYH, FANCD2, NBN, PALB2, and BRCA1 are associated with hereditary breast and ovarian cancer syndrome (HBOC), an autosomal dominant disorder typically manifesting around the age 30 and above (69). Some of these pathogenic variants can sometimes lead to conditions that manifest later in life. Some variants identified in this study were associated with familial and infantile disorders. However, the individuals carrying these variants remained healthy into adulthood. For example, the pathogenic variants *LZTR1:c.214dup*, *LZTR1:c.1785+1G>C*, and the likely pathogenic variant *LZTR1:c.509+3G>C*, along with *GJB2:c.505T>C*, associated with Noonan syndrome, were observed in Indian individuals who did not show signs of the associated disorders. Noonan syndrome is generally considered an autosomal dominant condition, but recent studies suggest that the *LZTR1* gene may be implicated in both dominant and recessive forms of the disorder (70,71). Similarly, Waardenburg syndrome (EDNRB), and *Van der* Woude syndrome 1 (IRF6) have also been reported autosomal dominant modes. This highlights that variants which are pathogenic in one context may be benign in another due to the interplay of genetic and environmental factors (72). Since the participants in the IndiGenomes study were healthy individuals, these variants might represent benign polymorphisms or variants with uncertain clinical significance in them, suggesting that they do not directly contribute to the disease phenotype in this particular population. In the Indian context, these same mutations did not appear to affect healthy individuals, which is consistent with findings from other studies. This discrepancy highlights the varying effects of genetic variants across populations due to differences in allele frequencies, genetic backgrounds, and environmental factors. In addition to these, several annotated pathogenic variants associated with dominant (Table 4) and recessive (Table 5) traits were identified, along with a substantial number of other clinically significant variants (SupplementaryData_S1.xlsx) in the IndiGenome dataset. In these cases, the identified variants may not cause disorders due to their insufficient disruptive effect or weak dominant influence. Studies reveal individuals with identified pathogenic variants who appear phenotypically normal, potentially due to factors such as gene dosage sensitivity (73), the functional impact of mutations (74,75), heterozygous carrier status (76), and other genetic modifiers (77). Findings in IndiGenomes highlight the critical role of population-specific genomic data in understanding rare genetic diseases (RGDs), as prevalence and mutation spectra can vary across ethnic groups (78).

Like other platforms and resources, GenTIGS has its limitations despite being a significant advancement in rare disease research. Regular updates are crucial to keeping the database current as new genetic disorders and variants are discovered. The integration of multi-omics data (such as transcriptomics and proteomics) would offer a deeper understanding of disease mechanisms. Additionally, expanding the representation of clinical case studies, particularly from diverse populations like India, and incorporating data from other regions and ethnic groups, would improve genotype-phenotype correlations and enhance its global relevance in rare genetic disorder research.

### Conclusion

GenTIGS is a powerful and versatile platform for RGDs research and clinical genomics. By integrating detailed genetic and clinical data with advanced analytical tools, it deepens our understanding of the genetic basis of diseases, particularly in the Indian context. Its alignment with global databases enhances its reliability, and features like disorder prediction and variant annotation show promise for improving diagnosis and treatment of RGDs. The expansion of the platform to analyse population-wide genomic data, advancing precision medicine for RGDs. Genetic variant pathogenicity is influenced by population-specific factors, such as allele frequencies and environmental conditions. A variant benign in one population may be pathogenic in another, emphasizing the importance of tailored genetic assessments. Our comparative analysis indicates that variants identified in Indian genomes may not have the same pathogenic impact as those found globally, reinforcing the need for region-specific genetic data. Understanding these population-specific differences is crucial for accurate diagnosis and personalized treatment strategies. By focusing on diverse populations, we can improve the identification of relevant genetic markers. GenTIGS, though a significant advancement in rare disease research, has limitations that require regular updates and the integration of multi-omics data for deeper insights. Expanding clinical case studies, particularly from diverse populations like India, would improve genotype-phenotype correlations and enhance its global relevance. GenTIGS may be a special relevance and impact in Southeast Asia and the Indian subcontinent, where genetic factors are likely to have a stronger correlation.

## Data Availability

All data produced are available in the manuscript and online at
https://db.tigs.res.in/gentigs/

## Acknowledgements

This research was supported by the Tata Institute for Genetics and Society (TIGS), India. Additionally, we extend our sincere appreciation to Dr. Surabhi Srivastva for her insightful suggestions and feedback provided on multiple occasions for the quality improvement and refining key aspects of this work.

## Author contributions

I.R. and R.M. conceived the study and the overall conceptual design. I.R. collected and compiled the data and developed programs for designing and creating the database and application modules used for browsing and analyzing the information. PS collected and curated the data and tested the workflow of GenTIGS. S.M. tested the workflow model and assisted in data collection. I.R. and S.M. drafted the manuscript. All authors have read and approved the manuscript.

## Competing interests

The authors declare no competing interests.

## Financial Support

Tata Institute for Genetics and Society (TIGS) fully funds this project.

## Supplemental data

The supplemental data described in the manuscript can be downloaded from https://db.tigs.res.in/gentigs/download/.

## Notes

### Competing Interest Statement

The authors have declared no competing interest.

### Funding Statement

The Tata Institute for Genetics and Society (TIGS) provides full financial support for this project, which aims to bridge the gap between genomic data and patient care, thereby improving information accessibility.

### Author Declarations

The study exclusively utilized publicly available human data, sourced from the following platforms: PubMed: Initial information on rare genetic disorders (RGDs). ClinVAR (https://www.ncbi.nlm.nih.gov/clinvar/) and ClinGen (https://clinicalgenome.org/): Genetic variants and associated genes for RGDs. OMIM (https://www.omim.org/): Data on inheritance patterns and disorder categories. NCBI-MedGen (https://www.ncbi.nlm.nih.gov/medgen/): Clinical genetic information. GARD (https://rarediseases.info.nih.gov/): Clinical symptoms of RGDs. GTR (https://www.ncbi.nlm.nih.gov/gtr/): Disorder synonyms. NCBI-Gene (https://www.ncbi.nlm.nih.gov/): Gene information and annotations. Human Genome GRCh38.p14 (https://www.ncbi.nlm.nih.gov/): Genomic sequences. MalaCards (https://www.malacards.org/): Supplementary data and disorder subcategories. These databases were used to gather and organize comprehensive, up-to-date information on rare genetic disorders, ensuring the study's data was both thorough and current.

